# Persistent Neurological Symptoms After COVID-19 Lack Evidence of Adaptive CNS Immune Activation

**DOI:** 10.64898/2026.08.10.26359944

**Authors:** Deborah K. Erhart, Luisa T. Balz, Ioanna Giotaki, Lynn Matits, Rüdiger Groß, Franziska Bachhuber, Jan Münch, Iris-Tatjana Kolassa, Dirk Fitzner, Ingo Uttner, Dorothée Lulé, Jan Lewerenz, Peter Lange, Hayrettin Tumani

## Abstract

Persistent neurological symptoms are among the most disabling manifestations of post-COVID-19 syndrome (PCS), yet the contribution of ongoing CNS immune activation remains uncertain. CSF studies including clinically relevant COVID-19 recovered control cohorts are scarce. In this prospective single-center study, we enrolled 50 patients fulfilling the WHO criteria for PCS (COVID^post^, mean age ± standard deviation [SD] 43.41 ± 11.99 years, 30 % male, 70 % female) and 50 individuals who had fully recovered from COVID-19 (COVID^reco^, mean age ± SD 39.38 ± 13.45, 42 % male, 58 % female). Both cohorts were comparable regarding age (*p* = 0.07), sex (*p* = 0.30), and education (*p* = 0.84). All participants underwent paired CSF and serum analyses together with comprehensive neuropsychological assessment. Routine CSF parameters, blood-CSF barrier integrity, oligoclonal bands (OCB), SARS-CoV-2 RNA in CSF and blood, pathogen-specific antibody indices, and neuronal autoantibodies were investigated. Despite marked differences in cognitive performance (*p* < 0.001) and fatigue severity (*p* < 0.001), patients with PCS showed no evidence of disease-specific CSF abnormalities compared to recovered controls. Routine CSF parameters, blood-CSF barrier dysfunction, CSF-restricted OCB, SARS-CoV-2 RNA in CSF and blood, intrathecal SARS-CoV-2 antibody synthesis, polyspecific antiviral immune responses, and neuronal autoantibodies were comparable between groups. SARS-CoV-2-specific IgG concentrations in CSF correlated positively with serum concentrations (COVID^post^: *r* [95%CI] = 0.78 [0.62 - 0.87]; COVID^reco^: *r* [95%CI] = 0.86 [0.75 - 0.92]; both *p* < 0.001) and albumin quotient (COVID^post^: *r* [95%CI] = 0.52 [0.26 - 0.71], *p* < 0.001; COVID^reco^: *r* [95%CI] = 0.37 [0.10 - 0.60]; *p* = 0.01), consistent with passive transfer across the blood-CSF barrier rather than compartmentalized intrathecal immune activation. Furthermore, SARS-CoV-2- specific antibody measures were not associated with cognitive performance (*p* > 0.72) or fatigue severity (*p* > 0.88). This study provides no evidence that persistent neurological symptoms after COVID-19 are accompanied by ongoing adaptive CNS immune activation, disease-specific neuronal autoimmunity, or intrathecal SARS-CoV- 2-specific humoral immune responses. The inclusion of a carefully phenotyped COVID-19 recovered comparison cohort strengthens the conclusion that routine CSF abnormalities largely do not seem to reflect mechanisms specific to PCS. These findings argue against routine CSF diagnostics as a source of disease-specific biomarkers in unselected PCS patients and support future studies focusing on alternative mechanisms underlying persistent neurological symptoms.

## Introduction

Since the emergence of the severe acute respiratory syndrome coronavirus 2 (SARS-CoV-2), increasing attention has been directed toward the long-term condition following acute coronavirus disease 2019 (COVID-19).^1^ While many individuals recover completely, a substantial proportion either experience persistent or develop new neurological, mainly cognitive, and other systemic symptoms. These symptoms can last for months to years beyond the acute phase of four weeks after initial symptoms of COVID-19 and are commonly referred to as post-COVID-19 syndrome (PCS).^2,3^ Approximately 6% to 15% of individuals infected with SARS-CoV-2 report persistent post-acute, predominantly neuropsychiatric, symptoms.^2–4^ Frequently reported manifestations include fatigue/reduced physical performance (incl. post-exertional malaise), cognitive impairment, headache, disturbances of taste/smell, various pain syndromes, and sleep disorders, suggesting potential ongoing alterations within the CNS and immune response.^1,4,5^ While a vast proportion of patients recovers slowly, up to 4% of post-COVID-19 patients still complain about persistent symptoms after two years, with substantial impact on patient’s quality of life and socioeconomic burden.^6^ A comprehensive understanding of PCS specific immune response profile in association with well-defined clinical characteristics is essential to define potential therapeutic targets.

CSF analysis provides a valuable approach for investigating direct neuroinvasive involvement and CNS immune processes, including intrathecal immunoglobulin synthesis and neuronal autoantibody responses. A substantial proportion of studies and case series investigating neurotropism of SARS-CoV-2 in acute COVID-19 with neurological manifestation could not detect SARS-CoV-2 RNA in CSF via rRT-PCR.^7–13^ Yet, there are case reports and case series from the early period of the COVID-19 pandemic describing positive SARS-CoV-2 RNA results in patients with (meningo- )encephalitis/myelitis (summarized in Luis et al^14^). Additionally, virus-specific RNA and transcripts of SARS-CoV-2 related proteins, e.g. nucleocapsid (N) and trimeric spike (S1) protein as well as viral entry receptors angiotensin converting enzyme 2 (ACE2), and transmembrane protease serine subtype 2 (TMPRSS2) were found in post-mortem analyses along the olfactory tract (bulbus and mucosa) as well as in cells of the choroid plexus and ependymal cells of the ventricles, brainstem, and cranial nerves, all in close relationship to the CSF space.^15–18^

The number of comprehensive studies combining CSF and serum analyses in PCS investigating SARS-CoV-2 specific and polyclonal immunity as well as COVID-19-related autoimmunity of the CNS is limited so far.^10,19–21^ Moreover, a vast majority of previous studies focused solely on patients with PCS and lacked the clinically relevant comparison cohort of individuals who had fully recovered from acute COVID-19.^10,19,21^ Moreover, immune dysregulation may contribute to the pathophysiology of persistent symptoms after COVID-19.^22–24^ Both systemic and CNS immune activation have been proposed as possible mechanisms underlying prolonged neurological complaints.^25,26^ In particular, activation via characteristic morphological changes of microglia in hypothalamus,^27^ CNS autoimmune responses,^21^ cytokine dysregulation,^28,29^ the persistence of pro-inflammatory transcription status of circulating B, T, and natural killer (NK) cells, and monocytes,^29,30^ dysfunctions in circulating immune cells, e.g., CD4+ and CD8+ (SARS-CoV-2 specific) T cells, even in CSF,^24,31^ and increased T cell activity in the brainstem and spinal cord^32^ have been discussed as contributors to post-infectious neuroinflammation. However, the extent to which intrathecal immune activation differs between individuals with and without persisting symptoms after COVID-19 remains insufficiently understood.

This study therefore investigates markers of intrathecal immune activation in individuals after COVID-19 infection, comparing participants with persistent symptoms (COVID^post^) to those without ongoing complaints (COVID^reco^). To the best of our knowledge, this is the first study incorporating a COVID-19 fully recovered control cohort incl. their combined CSF and serum analyses and group allocation being based on both self-reported complaints and the results of a comprehensive neuropsychological assessment. By examining immunological alterations within the central nervous system, this work aims to contribute to a better understanding of the mechanisms underlying post-COVID-19 manifestations and their potential neuroimmunological basis. To address this, we investigated (I) if there is any evidence of blood-CSF barrier dysfunction in the two cohorts COVID^post^ and COVID^reco^ as was recently reported for acute COVID-19,^7,8,33^ (II) if there is any evidence of acute (CSF white blood cell count [WBCC] and lactate) or chronic intrathecal immune activity according to CSF-specific oligoclonal bands, polyspecific immune activity or intrathecal synthesis of SARS-CoV-2 antibodies, (III) if there is any evidence of SARS-CoV-2 RNA in CSF or blood, (IV) if any anti-neuronal antibodies or surface antibodies can be detected in CSF and serum in the two cohorts, and (V) if any CSF or serum findings are associated with cognitive impairment and/or fatigue status.

## Materials and methods

### Study design and participants

All participants were recruited prospectively from the post-COVID-19 outpatient unit of the Department of Neurology, Ulm University Hospital, Germany. In our cross-sectional, monocentric study, 50 patients with PCS (COVID^post^), diagnosed according to the clinical case definition of the WHO Delphi consensus criteria^34^ by experienced physicians in the field of post-viral syndromes (DKE and HT), were included from March 2023 to November 2024. As illustrated in the flow diagram (Fig. 1), 11 patients who had also presented to the post-COVID-19 outpatient unit were excluded prior to study inclusion based on the following diagnoses established during the extended diagnostic workup: Alzheimer’s disease (dementia *n* = 2, mild cognitive impairment (MCI) *n* = 1), Waldenstrom’s macroglobulinemia *n* = 1, Sjoegren’s syndrome *n* = 3, multiple sclerosis *n* = 1, antibody-negative autoimmune encephalitis *n* = 1, and depression, severe episode, *n* = 2. The extended diagnostic workup comprised an MRI of the brain, a comprehensive neuropsychological assessment, and a diagnostic lumbar and venipuncture. According to the clinical case definition,^34^ patients were seen at least 12 weeks after an acute COVID-19 infection and with at least one, in the case of our study, neurological symptom, that had been persisting since these 12 weeks following the acute infection. As PCS is a diagnosis of exclusion, only patients in whom alternative medical conditions or diagnoses that could account for the reported symptoms had been ruled out were eligible for inclusion. This process included a comprehensive internal medicine evaluation, e.g., endocrinology, cardiology, gastroenterology, and rheumatology, and cancer screening (age-appropriate). Furthermore, men underwent urological assessment and women gynecological evaluation.

**Fig. 1.**
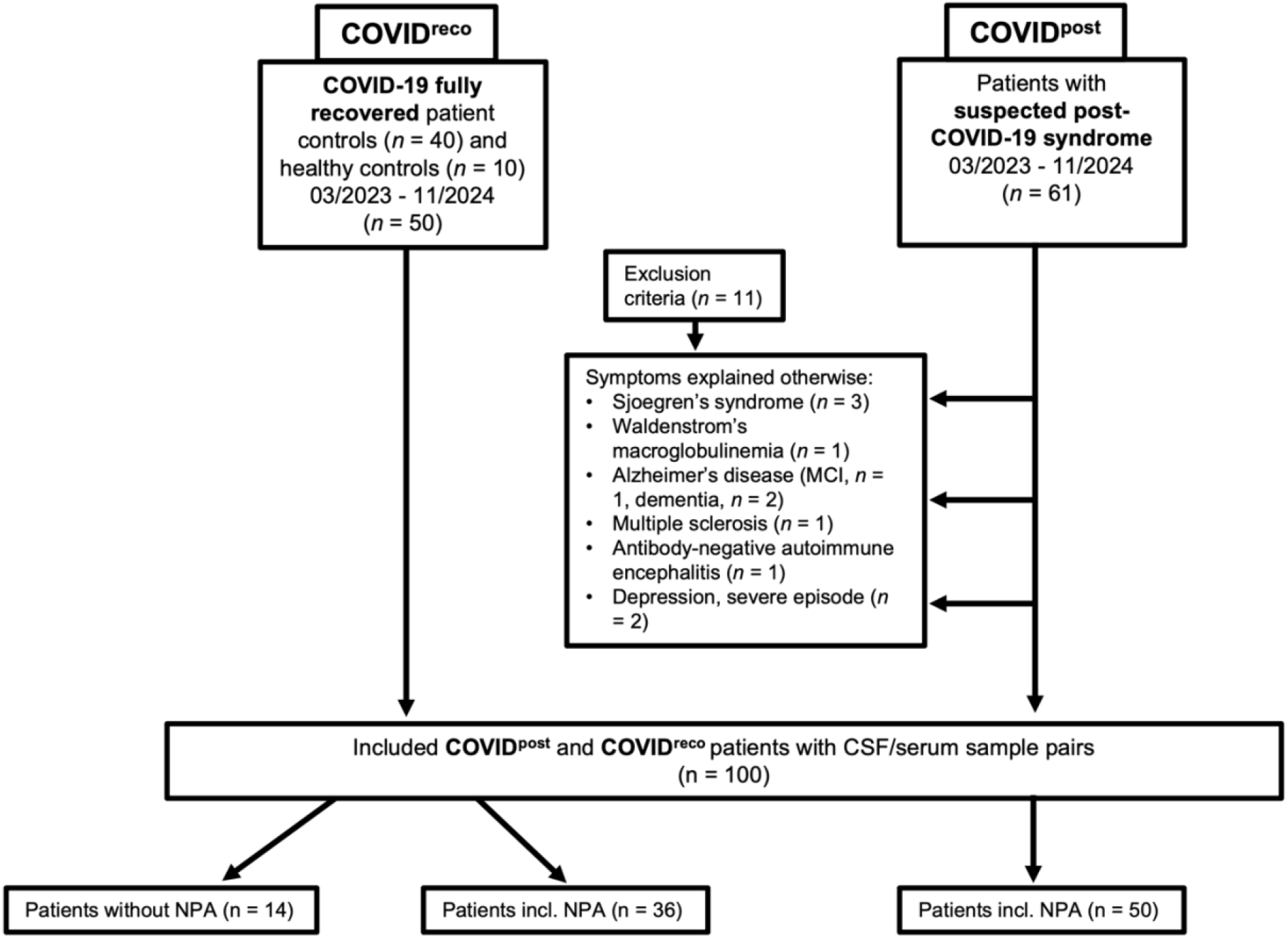
Patient recruiting process. COVID-19 fully recovered (COVID^reco^) and patients with suspected post-COVID-19 syndrome (PCS) (COVID^post^) were recruited in the Department of Neurology at Ulm University Hospital, Germany. The exclusion criteria for the COVID^post^ cohort are listed above.

Additionally, we included 50 patients who also had COVID-19 infection in the medical history but without any persistent symptoms since then (COVID^reco^). Patients of the COVID^reco^ cohort were either recruited in our emergency room or different outpatient units of the Ulm University Hospital (*n* = 40) who received at least a lumbar and venipuncture as part of their routine diagnostics and in whom neurodegenerative or neuroinflammatory diseases of the central nervous system were additionally ruled out by MRI of the brain. Furthermore, ten healthy volunteers, who had also suffered COVID-19, consented to be part of the study and also received lumbar puncture after a brain MRI. These volunteers were also assigned to the COVID^reco^ cohort (total n = 50). Thirty-six participants of the COVID^reco^ cohort also underwent the same neuropsychological assessment as the COVID^post^ cohort. All included participants of both cohorts had a prior SARS-CoV-2 infection confirmed by either positive rRT-PCR testing or SARS-CoV-2 antigen rapid test. This study follows the STROBE guidelines.

### Neuropsychological assessment

The neuropsychological assessment (NPA) was conducted by one neuropsychologist (LTB). The examination included validated testing instruments for different cognitive domains. Attention (incompatibility, alertness, divided attention) was assessed using the German Testbatterie zur Aufmerksamkeitsprüfung (TAP).^35^ We evaluated working memory and verbal short-term memory with the Digit Span Test from the Wechsler Memory Scale-Revised (WMS-R) and non-verbal short-term and working memory with the Block-Tapping Test from WMS-R.^36^ The Rey-Osterrieth Complex Figure Test (ROCF) was employed for assessing non-verbal episodic memory^37^ and the Verbal Learning and Memory Test (VLMT) for verbal episodic memory.^38^ Verbal fluency was evaluated using the Regensburger Wortflüssigkeitstest (RWT),^39^ the German version of verbal fluency measures, and the Symbol Digit Modalities Test (SDMT) was used to measure selective and divided attention and information processing speed.^40^

Besides the neuropsychological examination, we also captured motor and cognitive fatigue with the 20-item questionnaire Fatigue Scale for Motor and Cognitive Functions (FSMC).^41^ Each item is rated on a five-point Likert scale ranging from one to five. The motor fatigue subscale comprises the following cut-offs: ≥ 22 mild fatigue, ≥ 27 moderate fatigue, and ≥ 32 severe fatigue. The cognitive fatigue score was categorized as follows: ≥ 22 mild fatigue, ≥ 28 moderate fatigue, and ≥ 34 severe fatigue. The total fatigue score is calculated by summing the scores of the cognitive and motor fatigue subscales. Cut-off values for the FSMC total score indicate mild fatigue at scores ≥ 43, moderate fatigue at scores ≥ 53, and severe fatigue at scores ≥ 63.^41^ Established age- and education-corrected z-scores are currently available only for the SDMT (Supplementary Table S1).^40^ The observed proportion of participants in our COVID^reco^ cohort with below-average^42^ SDMT performance (11.1%) was consistent with the expected base rate in healthy populations, supporting the validity of the control cohort as a reference population.^43,44^ Therefore, all 20 cognitive subtests of our neuropsychological assessment were z-standardized based on the data of the COVID^reco^ cohort using their pooled mean and standard deviation (SD) as formerly described.^45^ A global cognition composite score was built by averaging the respective z-scores of the subtests to obtain an overall measure of cognitive performance across multiple cognitive domains as described above and to reduce the influence of variability in individual test scores.

### CSF routine diagnostics

All study participants received lumbar (LP) and venipuncture (serum and EDTA-plasma) from the same neurologist (DKE). After drawing, CSF and serum samples were brought immediately (< 5 min) to the CSF and autoimmune laboratory of Ulm University Hospital. All samples were handled according to the guidelines of the German Society for Cerebrospinal Fluid Diagnostics and Clinical Neurochemistry (DGLN) and the German Society of Neurology (DGN).^46^ Native CSF was used for cell count (leukocytes/µl and erythrocytes/µl) which was performed manually in the Fuchs-Rosenthal chamber (leukocyte count reference range: ≤ 4 leucocytes/µl; erythrocyte count reference range: 0 erythrocytes/µl). The remaining CSF and serum tubes were centrifuged at 2000 x *g* for 10 min (room temperature) and the supernatant was used for further (protein) analyses. Additionally, several aliquots were stored in our biobank at -80°C. For differential cell count analysis, the cell pellet was transferred to a Cellspin I cytocentrifuge (THARMAC GmbH, Waldsolms, Germany) and centrifuged at 1000 x *g* for 10 min. Cytospin preparations were subsequently stained according to the Pappenheim method. Differential cell counts were determined by manually classifying 100 cells under light microscopy, and the relative proportions of the individual cell populations were expressed as percentages.

Lactate in CSF was detected using a photometer (reference range: 1.7 - 2.6 mmol/l, AU400/AU680 Clinical Chemistry Analyzers [Olympus/Beckman Coulter, Krefeld, Germany]). A nephelometer (Atellica® NEPH 630 System [Siemens Healthcare GmbH, Erlangen, Germany]) was used to measure albumin (mg/l), and immunoglobulins A, M, and G (mg/l) in CSF and serum. Total protein (g/l) concentrations were determined using an immunoturbidimetric assay (Cobas c 503, Roche Diagnostics, Mannheim, Germany). We used CSF/serum albumin ratio (Q_Alb_) to describe the assessment of the blood–CSF barrier integrity. The upper limit (Q_Alb_lim_) was calculated as an age-dependent cut-off using the following formula: Q_Alb_lim_ < (4 + age/15) × 10^−3^).^47^ The following cut-offs were adopted for blood-CSF barrier dysfunction for values > Q_Alb_lim_: slight ≤ 10 × 10^−3^, moderate ≤ 20 × 10^−3^, and severe > 20 × 10^−3^.^48^

### Investigations of humoral immune response and antibody indices

Isoelectric focusing (IEF) was used to detect and count oligoclonal IgG bands (OCB) in CSF and serum as previously described.^49^ Quantitative intrathecal synthesis of total IgG, IgA, and IgM (Ig_Syn_) was calculated based on respective CSF/serum ratios for IgG, IgA, and IgM (Q_IgG_total_, Q_IgA_total_, Q_IgM_total_) relative to their respective upper reference limits (URL).^50^ URL for each Ig_CSF_/Ig_serum_ ratio was calculated using the Reiber formulas and defined as Q_lim_high_.^50^ A quantitative Ig_Syn_ is defined as a concentration of the respective immunoglobulin far exceeding the expected concentration by taking its serum concentration and the individual Q_Alb_ into account, i.e. a quantitative Ig_Syn_ was assumed for Q_IgG_total_, Q_IgA_total_, Q_IgM_total_ > Q_lim_high_.^50^ Furthermore, we also assessed pathogen-specific antibody indices (AI). First, pathogen-specific IgG levels against measles virus (M), rubella virus (R), varicella-zoster virus (Z), herpes-simplex virus type 1 (HSV1), Epstein-Barr virus (EBV), Cytomegaly virus (CMV), and the trimeric spike protein subunit S1 of SARS-CoV-2 (CoV) were measured in CSF and serum using an enzyme-linked immunosorbent assay (ELISA) according to the manufacturer’s instructions (Gold Standard Diagnostics Europe, formerly Genzyme Virotech; Rüsselsheim, Germany for M, R, Z, HSV1, EBV, and CMV; Virion\Serion, Würzburg, Germany for CoV). Accordingly, pathogen-specific CSF/serum IgG ratios (Q_spec_) and AIs were calculated by dividing Q_spec_ by Q_IgG_total_ for Q_IgG_total_ ≤ Q_lim_high_ and Q_spec_/Q_lim_high_ for Q_IgG_total_ > Q_lim_high_.^51^ For AI levels ≥ 1.5, we assumed pathogen-specific intrathecal synthesis of specific IgG.^51^

### SARS-CoV-2 PCR in CSF and blood

Viral RNA was isolated from 17.5 μl CSF and EDTA-plasma of the frozen (-80°C) aliquoted samples from the biobank using the Qiagen Viral RNA Mini Kit #52906 (Qiagen, Venlo, The Netherlands) according to the manufacturer’s instructions. Centrifugation was performed at 8000 x g, with the exception of the final wash step, which was performed at 20000 x g. Samples were mixed with lysis buffer (AVL) and incubated 20 min at room temperature. Samples were spiked with 5.6 μg carrier RNA, followed by vortexing and an additional 10 min incubation at room temperature. 560 μl ethanol was then added, samples were vortexed and briefly centrifuged to remove droplets from the lid, and the entire volume was then stepwise loaded onto columns. All subsequent steps were performed as instructed by the manufacturer. Viral RNA was eluted in 60 μl AVE buffer. RT-PCR for SARS-CoV-2-N and -ORF1b-nsp14 was performed using TaqMan Fast Virus 1-Step as previously described.^52^ All samples were measured in duplicates, values > cycle threshold 35 were defined as “not detected”.

### CNS autoantibody testing

As part of the routine diagnostics, serum samples (dilution 1:101) were tested using a commercial line blot for intracellular antigens (PNS 12 Ag, EUROLINE, Euroimmun, Lübeck, Germany) for anti-amphiphysin, anti-CV2, anti-PNMA2 (paraneoplastic Ma-2 antigen, anti-Ma-2/Ta), anti-Ri, anti-Yo, anti-Hu, anti-recoverin, anti-SOX1 (SRY-box transcription factor 1), anti-Titin, anti-Zic4 (Zic family member 4), anti-GAD65 (glutamic acid decarboxylase 65), and anti-Tr/DNER (Delta and Notch-like epidermal growth factor-related receptor). Simultaneously to the line blot, undiluted CSF and serum (dilution 1:50) were analyzed using indirect immunofluorescence on a commercial tissue-based assay (Neurologie-Mosaik 8, Euroimmun, Lübeck, Germany) made of monkey cerebellar, nerval, pancreatic and intestine sections for detecting: Anti-GAD65, anti-Yo, anti-Hu, anti-Ri, anti-CV2, anti-Ma, anti-amphiphysin.

Regarding CSF and serum analytes for anti-surface-receptor antibodies, a two-step procedure was performed. First, fixed rat brain sections (sagittal, coronar, axial) were treated separately with CSF (dilution 1:4) and serum (dilution 1:200) according to the protocol published by Dalmau et al.^53^ to evaluate the respective fluorescence intensity for the antibodies of the following receptors: Neuropil, NMDA, AMPA, GABAB, CASPR2 (contactin-associated protein-like 2), LGI1 (Leucine-rich, glioma inactivated protein 1), DPPX (dipeptidyl-peptidase-like protein-6), IgLON5 (IgLON family member 5). We graded negative (lack of any specific fluorescence) signal from weakly positive, moderately positive, and strongly positive intensities using a predefined grayscale template. Additionally, a cell-based indirect immunofluorescence assay (CBA/IIFT) for the anti-NMDA-R antibodies for CSF and serum was applied (Euroimmun, Lübeck, Germany). Undiluted CSF and serum (dilution 1:10) of those samples with any positive neuropil result on the unfixed rat brain sections were tested on a second commercial CBA/IIFT (Autoimmune-Enzephalitis-Mosaik 6, Euroimmun, Lübeck, Germany). The assay detected the following CNS-autoantibodies: anti-NMDA-R, anti-AMPA1-R, anti-AMPA2-R, anti-CASPR2, anti-LGI1-R, anti-GABA-B1-R, and anti-DPPX-R. For the anti-IgLON5-R, we used a separate CBA/IIFT (Euroimmun, Lübeck, Germany). All commercial assays were performed according to the manufacturer’s instructions and laboratory personnel was blinded to cohort assignment.

### Ethics

Written informed consent was obtained from each patient. This study was conducted under the principles of the Declaration of Helsinki and was approved by the ethics review committee of Ulm University (approval numbers 16/23 from 16^th^ of March 2023 and 20/10 from 20^th^ of May 2010).

### Statistical analysis

Statistical analyses were performed using R version 4.5.1 (R Core Team, 2025). Figures were generated using GraphPad Prism V.10.2.2 (GraphPad Software, La Jolla, CA) and R for macOS. Given the lack of prior effect size estimates, no formal a priori sample size calculation was performed. With 50 COVID^post^ patients and 50 COVID^reco^ patients, the study had approximately 70 % power to detect a medium effect size (*d* = 0.5) at a two-sided α of 0.05, allowing detection of moderate-to-large effects while smaller differences may have remained undetected (G*Power V.3.1.9.6).^54^ The two cohorts (COVID^post^ and COVID^reco^) were dichotomized according to the presence of PCS according to the WHO Delphi consensus criteria of the clinical case definition.^34^ Categorical variables are given as absolute and relative frequencies. Continuous variables are presented as means (mean) and standard deviations (SD).

Assumptions were assessed visually using the *check_model* function from the performance package.^55^ Due to the presence of influential cases, robust Yuen’s *t*-tests based on trimmed means (*tr* = 0.2; WRS2 package)^56^, with bootstrapped confidence intervals, were used to assess group differences between 50 COVID^post^ patients and 50 COVID^reco^ patients in demographic data, cognition, fatigue, and CSF routine findings. Differences between groups for categorical variables were evaluated using Fisher’s exact tests (including Odds ratio (OR)) and χ² tests otherwise. Between-group comparisons of AIs were performed using the Brunner-Munzel test, a robust nonparametric test that does not assume homogeneity of variances. Correlation was assessed using the robust (regarding outliers), non-parametric percentage bend correlation with 95% CI of the correlation coefficient *r*. To examine whether the associations between SARS-CoV-2-specific antibody measures and neuropsychological outcomes differed between cohorts, linear regression models including interaction terms between group status and the respective antibody measures were conducted. Heteroscedasticity-robust standard errors and bootstrapped confidence intervals were used. Continuous predictors were z-standardized prior to analysis, and group membership was effect-coded. Effect sizes were quantified using partial eta-squared (η²p). For all analyses, an α-level of 0.05 was considered statistically significant.

## Results

### Demographic and clinical data of the study population

The two clinical cohorts COVID^post^ and COVID^reco^ were comparable with regard to age, sex, COVID-19 vaccination status, education, and time since acute COVID-19 to LP and NPA, respectively, with no significant between-group differences observed (Table 1). Based on the WHO Clinical Progression Scale for acute COVID-19,^57^ all participants were classified as state 1 (ambulatory mild disease), except for two individuals in the COVID^post^ cohort who met criteria for state 2 disease (hospitalized: moderate disease), with no difference between cohorts (*p* = 0.51). The mean time ± standard deviation [SD] from acute COVID-19 to the first PCS symptoms was 7.3 ± 18.3 days. Cognitive function and fatigue according to the FSMC was worse in the COVID^post^ cohort than in the COVID^reco^ cohort (Table 1). Twenty-three (46 %) COVID^post^ patients showed cognitive function below average^42^ according to the global cognition composite score consistent with cognitive impairment in contrast to two participants (6 %) of the COVID^reco^ cohort (*p* < 0.001, OR [95%CI] = 0.07 [0.01 - 0.33]). Additionally, fatigue severity according to the FSMC differed between cohorts (*p* < 0.001). Nearly all COVID^post^ patients (48/50; Supplementary Table S1) exhibited severe fatigue (96 %), whereas the majority of recovered individuals reported no fatigue (23/36, 64 %) or only mild fatigue (10/36, 28 %). Severe fatigue, not associated with PCS, was observed in only one recovered participant (3 %). Notably, the COVID^post^ cohort exhibited a higher prevalence (21/50, 42 %) of psychiatric lifetime diagnoses (*p* = 0.004), predominantly depression (16/50, 32 %).

**Table 1.** Demographic, clinical, and neuropsychological data of the COVIDpost and COVIDreco cohorts.

| Parameter | COVID<sup>post</sup><br>(*n* = 50) | COVID<sup>reco</sup><br>(*n* = 36) | Test statistics<sup>a</sup> |
| --- | --- | --- | --- |
| Age (mean ± SD, years) | 43.41 ± 11.99 | 39.38 ± 13.45 | $t = 2.03$, $p = 0.07$ |
| Sex | | | $\chi^2(1) = 1.09$, $p = 0.30$ |
| male *n*, % | 15 (30 %) | 21 (42 %) |  |
| female n, % | 35 (70 %) | 29 (58 %) |  |
| COVID-19 vaccinated (x/total), % | 46/50; 92 % | 36/36; 100 % | $\chi^2(1) = 0.01, p = 0.91$ |
| Hospitalization during COVID-19 (x/total), % | 2/50; 4 % | 0/36 | $p = 0.50$ |
| Education (mean ± SD, years) | 14.49 ± 1.95 | 14.35 ± 2.88 | $t = -0.21, p = 0.84$ |
| Time since COVID-19 to LP (mean ± SD, years) | 1.85 ± 0.88 | 1.49 ± 0.96 | $t = 1.60, p = 0.11$ |
| Time since COVID-19 to NPA (mean ± SD, years) | 1.85 ± 0.88 | 1.52 ± 0.96 | $t = 1.37, p = 0.19$ |
| Global cognition (mean ± SD, composite score) | -1.28 ± 1.76 | 0.00 ± 0.49 | **$t = -4.3, p < 0.001$** |
| FSMC total (mean ± SD) | 84.80 ± 9.31 | 34.17 ± 14.06 | **$t = 16.47, p < 0.001$** |
| FSMC cognition (mean ± SD) | 42.62 ± 5.30 | 17.61 ± 7.78 | **$t = 15.04, p < 0.001$** |
| FSMC motoric (mean ± SD) | 41.98 ± 5.06 | 16.56 ± 6.65 | **$t = 17.23, p < 0.001$** |

Only patients from the COVID<sup>reco</sup> cohort with complete neuropsychological data were included in the analyses presented in this table ( $n = 36$ ). <sup>a</sup>P-values and test statistics for continuous variables were obtained using a bootstrapped Yuen's test (trimmed means, $tr = 0.2$ ). Fisher's exact test was used for categorical variables. P-values $< 0.05$ were considered statistically significant and highlighted in bold. Contin SD: standard deviation, NPA: neuropsychological assessment, FSMC: fatigue scale for motor and cognitive functions.

The most commonly reported post-infectious symptoms in the COVID ^post^ cohort (Fig. 2, Supplementary Table S2) were fatigue, including post-exertional malaise (PEM) in 100%, subjective cognitive deficits (100 %), sleep disorder (31/50, 62 %), headache (25/50, 50 %), and myalgia (24/50, 48 %).

**Fig. 2.**
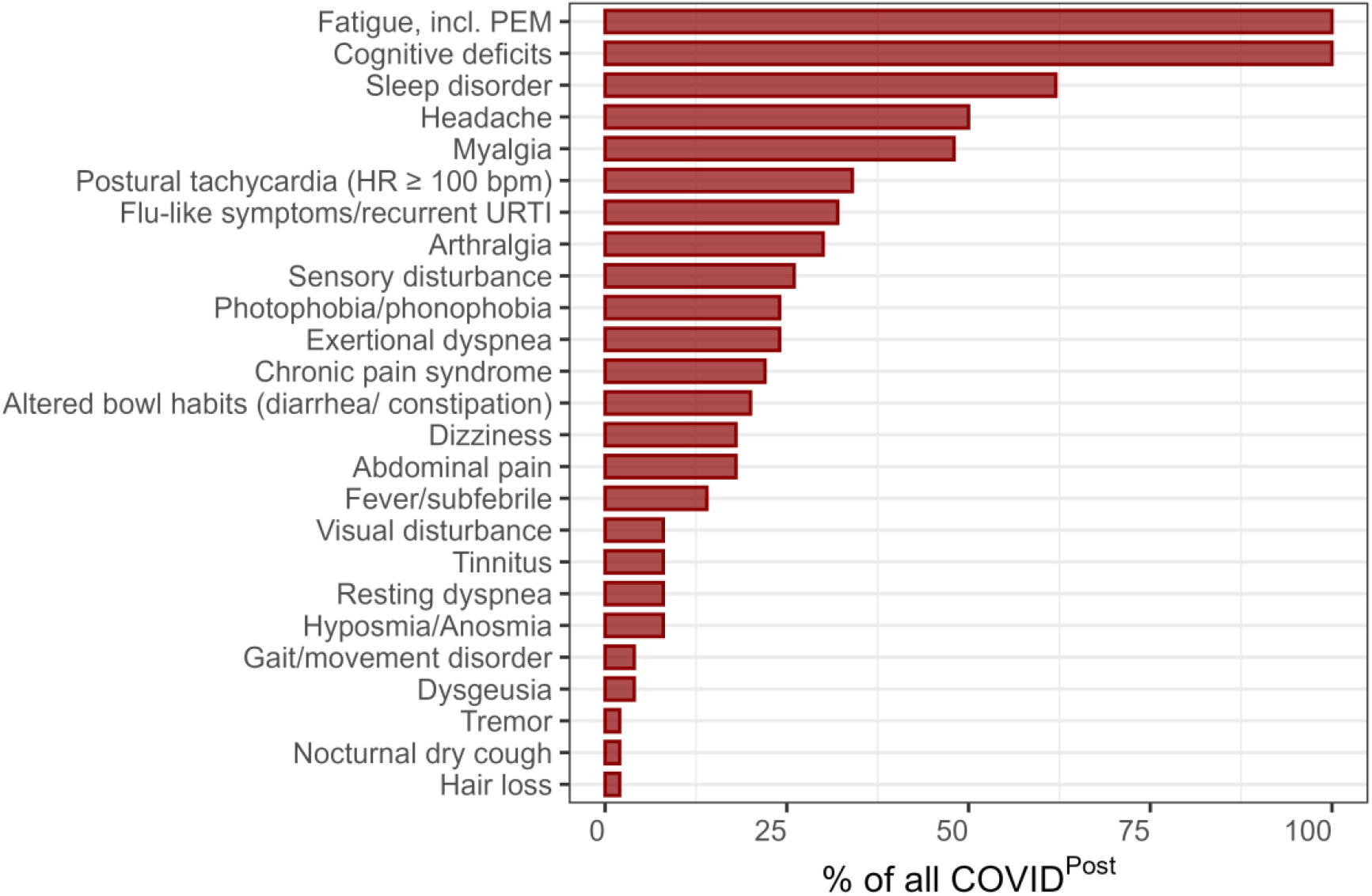
Overview of post-infectious symptoms in the COVID^post^ cohort. Values regarding each post-infectious symptom are given as relative frequencies of the 50 COVID^post^ patients. PEM: post-exertional malaise, HR: heart rate, bpm: beats per minute, URTI: upper respiratory tract infection.

### CSF routine findings

Overall, the two study cohorts COVID^post^ and COVID^reco^ were comparable regarding the investigated CSF routine parameters leukocyte count, total protein, lactate, Q_Alb_, Q_IgG_total_, and CSF-specific OCB (Table 3, Supplementary Table S3). In the COVID^post^ cohort, 6/50 (12%) patients had lympho-monocytic pleocytosis (leukocyte count ≥ 5/µl, 64 - 88 % lymphocytes (range) and 10 - 15 % monocytes (range)) with 2 - 14% activated lymphocytes (range) and 4% activated monocytes. In COVID^reco^ only one participant presented with pleocytosis (5 CSF leucocytes/µl) and no signs of activation. The distribution of OCB patterns did not differ between the two cohorts (*p* = 0.331). Six patients from the COVID^post^ cohort (12 %) and two participants of the COVID^reco^ cohort (4 %) had CSF-specific OCB with no difference observed (*p* = 0.27). Only 3/7 patients with elevated leukocyte count, all of the COVID^post^ cohort, had CSF-specific OCBs. None of the participants exhibited a Q_IgG_total_ exceeding Q_lim_high_.

**Table 3.** CSF routine findings in the whole COVID^reco^ and COVID^post^ cohort.

| Parameter | COVID<sup>post</sup> | COVID<sup>reco</sup> | Test statistics<sup>a</sup> |
| --- | --- | --- | --- |

|  | (n = 50) | (n = 50) |  |
| --- | --- | --- | --- |
| **Leukocytes/μl (mean ± SD)** | 2.18 ± 2.75 | 1.64 ± 1.31 | $t = 0.16, p = 0.86$ |
| **Total protein [mg/l] (mean ± SD)** | 366.12 ± 138.95) | 336.62 ± 124.95 | $t = 1.13, p = 0.25$ |
| **Lactate [mmol/l] (mean, SD)** | 1.47 ± 0.14) | 1.43 ± 0.17 | $t = 1.29, p = 0.21$ |
| **Q<sub>Alb</sub> [$\times 10^{-3}$] (mean, SD)** | 5.45 ± 2.13 | 5.08 ± 2.06 | $t = 1.09, p = 0.28$ |
| **Q<sub>IgG_total</sub> [$\times 10^3$] (mean, SD)** | 2.61 ± 1.10) | 2.25 ± 0.96) | $t = 1.09, p = 0.28,$ |
| **CSF-specific OCB (x/total), %** | 6/50, 12 % | 2/50, 4 % | OR [95%CI] = 3.24<br>[0.54; 34.43], $p = 0.27$ |

Means and standard deviations (SD) are reported for continuous variables. Categorical variables are presented as positive absolute and relative frequencies from total $n$ of each cohort. <sup>a</sup>P-values and test statistics for continuous variables were obtained using a bootstrapped Yuen's test (trimmed means, $tr = 0.2$ ). P-values < 0.05 were set as statistically significant. Fisher's exact test was used for the categorical variable CSF-specific OCB.

### SARS-CoV-2 RNA in CSF and blood

Reverse RT-PCR for SARS-CoV-2 RNA in CSF and blood was negative in all investigated samples of the COVID^post^ and COVID^reco^ cohort (*n* = 100).

### Blood-CSF barrier function in the two post-COVID-19 cohorts with and without persistent symptoms

In both groups, approximately every fifth participant (COVID^post^: 10/50 [20 %], COVID^reco^: 11/50 [22 %]) showed a Q_Alb_ above the upper reference indicative of blood-CSF barrier dysfunction (*p* = 0.59), which was usually slight (range: COVID^post^ 5.9 - 7.7 × 10^−3^, COVID^reco^ 6.2 - 9.4 × 10^−3^), except one patient from the COVID^reco^ cohort (10.6 × 10^−3^). No differences were observed between the COVID^post^ and COVID^reco^ cohorts with respect to SARS-CoV-2-specific IgG concentrations in either serum (mean ± SD: 84.39 ± 127.39 vs. 97.16 ± 87.29 U/ml; *p* = 0.08) or CSF (0.22 ± 0.28 vs. 0.53 ± 2.10 U/ml; *p* = 0.77).

The higher the serum SARS-CoV-2 IgG concentrations were (Fig. 3), the more SARS-CoV-2 IgG was found in the CSF in both COVID^post^ (r [95%CI] = 0.78 [0.62 - 0.87], *p* < 0.001) and COVID^reco^ (r [95%CI] = 0.86 [0.75 - 0.92], *p* < 0.001). In addition, CSF SARS-CoV-2-specific IgG levels correlated with Q_Alb_ in both the COVID^post^ cohort (*r* [95%CI] = 0.52 [0.26 - 0.71], *p* < 0.001) and the COVID^reco^ cohort (*r* [95%CI] = 0.37 [0.10 - 0.60], *p* = 0.01) reflecting the slight blood-CSF barrier dysfunction.

**Fig. 3.**
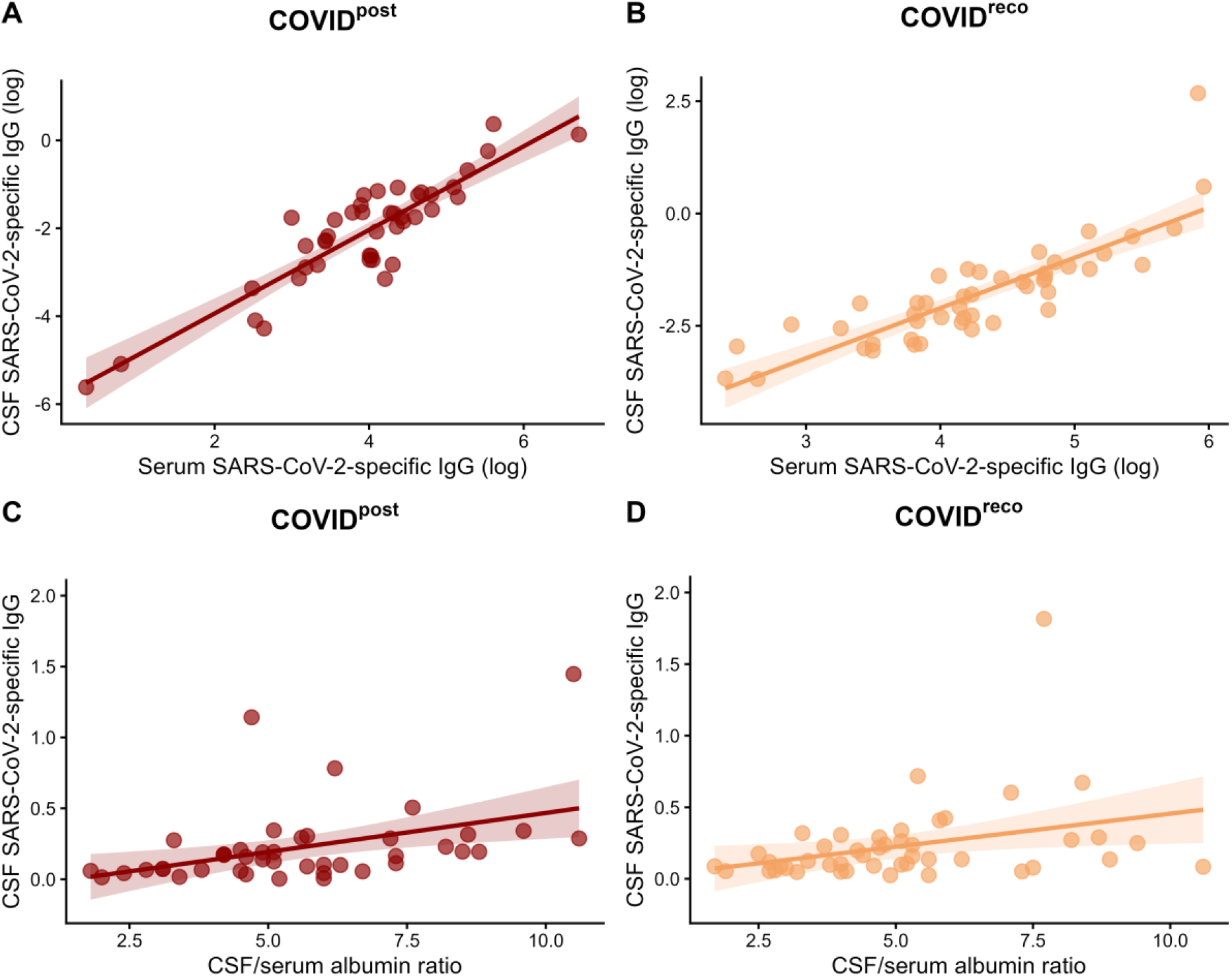
Association between Q_Alb_ and CSF and serum SARS-CoV-2-specific IgG concentrations. Scatterplot illustrating the association between ln-transformed CSF and serum SARS-CoV-2-specific IgG concentration [U/ml] in **A**) COVID^post^ and **B**) COVID^reco^ and between CSF SARS-CoV-2-specific IgG and Q_Alb_ in the two designated cohorts **C**) and **D**). The correlation was assessed using percentage bend correlation. Red points indicate COVID^post^ patients, while orange points indicate COVID^reco^ participants. For visualization purposes, the y-axis is restricted to values ≤ 2.0; one outlying observation in the COVID^reco^ group (SARS-CoV-2 IgG = 14.49 U/ml) is therefore not displayed.

### Investigations on polyspecific immune activation in PCS

Virus-specific AI analysis revealed elevated CoV-AIs (≥ 1.5) in only two participants, one from the COVID^post^ cohort (P1 in Figure 4) and one from the COVID^reco^ cohort (P5 in Fig. 4). The mean CoV-AI ± SD was 1.04 ± 0.51 in COVID^post^ patients and 1.40 ± 2.43 in fully recovered individuals. Additionally, CoV-AIs showed a trend toward higher values in the COVID^reco^ cohort; however, the between-group difference showed only a trend toward statistical significance (*p* = 0.053).

**Figure 4.**
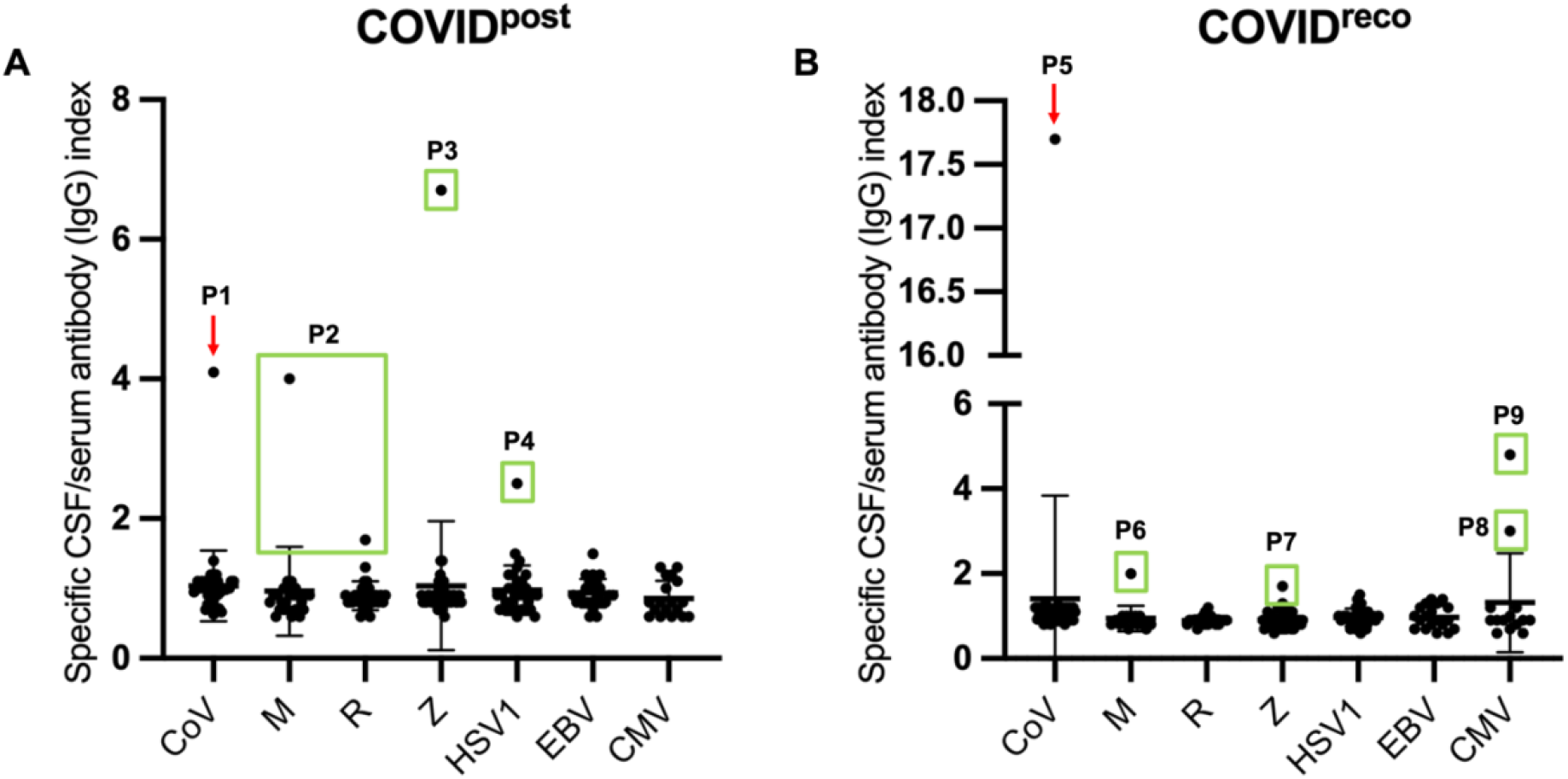
Different virus specific CSF/serum antibody (IgG) indices (AI) in the COVID^post^ and COVID^reco^ cohort. Scatterplots illustrating CSF/serum IgG AIs for different viruses: SARS-CoV-2 (CoV), measles (M), rubella (R), varicella-zoster (Z), herpes-simplex type 1 (HSV1), Epstein-Barr (EBV), and Cytomegaly virus (CMV) in the **A**) COVID^post^ and **B**) COVID^reco^ cohort. Data are shown as mean with standard deviation (SD). AIs were calculated using the Reiber formula.^51^ AI-levels ≥ 1.5 were considered elevated.^51^ Red arrows represent two patients (P1, P5) in the COVID^post^ cohort with elevated CoV-AI, and green boxes represent patients with elevated AIs against viral pathogens other than SARS-CoV-2 (P2-P4, P6-P9). For visualization purposes, the y-axis is divided in **B**.

Elevated virus-specific AIs were occasionally detected in both the COVID^post^ (four patients) and COVID^reco^ (five patients) cohorts (Table S4), including antibodies against measles virus (*n* = 2), rubella virus (*n* = 1), varicella-zoster virus (*n* = 2), HSV1 (*n* = 1), and CMV (*n* = 2). Patient 2 had an incomplete MRZ-reaction with CSF-specific OCB, but without any specific MRI findings regarding chronic inflammatory CNS disease.

### Neuronal surface and anti-neuronal antibody investigations related to PCS

The presence of neuronal surface antibodies in serum using commercial cell- and tissue-based assays (Table 4) was comparable (*p* = 0.99) in the COVID^post^ (4/50; 8%) and COVID^reco^ cohort (5/50; 10%). The detected antibodies comprised neuropil antibodies in seven cases (only in fixed rat brain section without otherwise confirmed) and one case each lower-titer CASPR2- and GABA-B-R antibodies (1:10) in serum (only in IIFT without confirmation in fixed rat brain sections). Notably, the participant positive for GABA-B-R antibodies additionally exhibited a pronounced immunofluorescence signal targeting Purkinje cells in the IIFT. And the other participant from the recovered COVID-19 cohort with serum CASPR2-antibodies suffered from idiopathic intracranial hypertension and was also tested positive for serum and CSF neuropil antibodies. However, this patient exhibited no symptoms of antibody-mediated encephalitis.^58^ Detailed CSF and brain MRI analysis showed no signs of intrathecal inflammation, as additionally reflected by a normal leukocyte count (1/µL), Q_Alb_ 2.8 × 10^−3^, and no CSF-specific OCB.

**Table 4.**
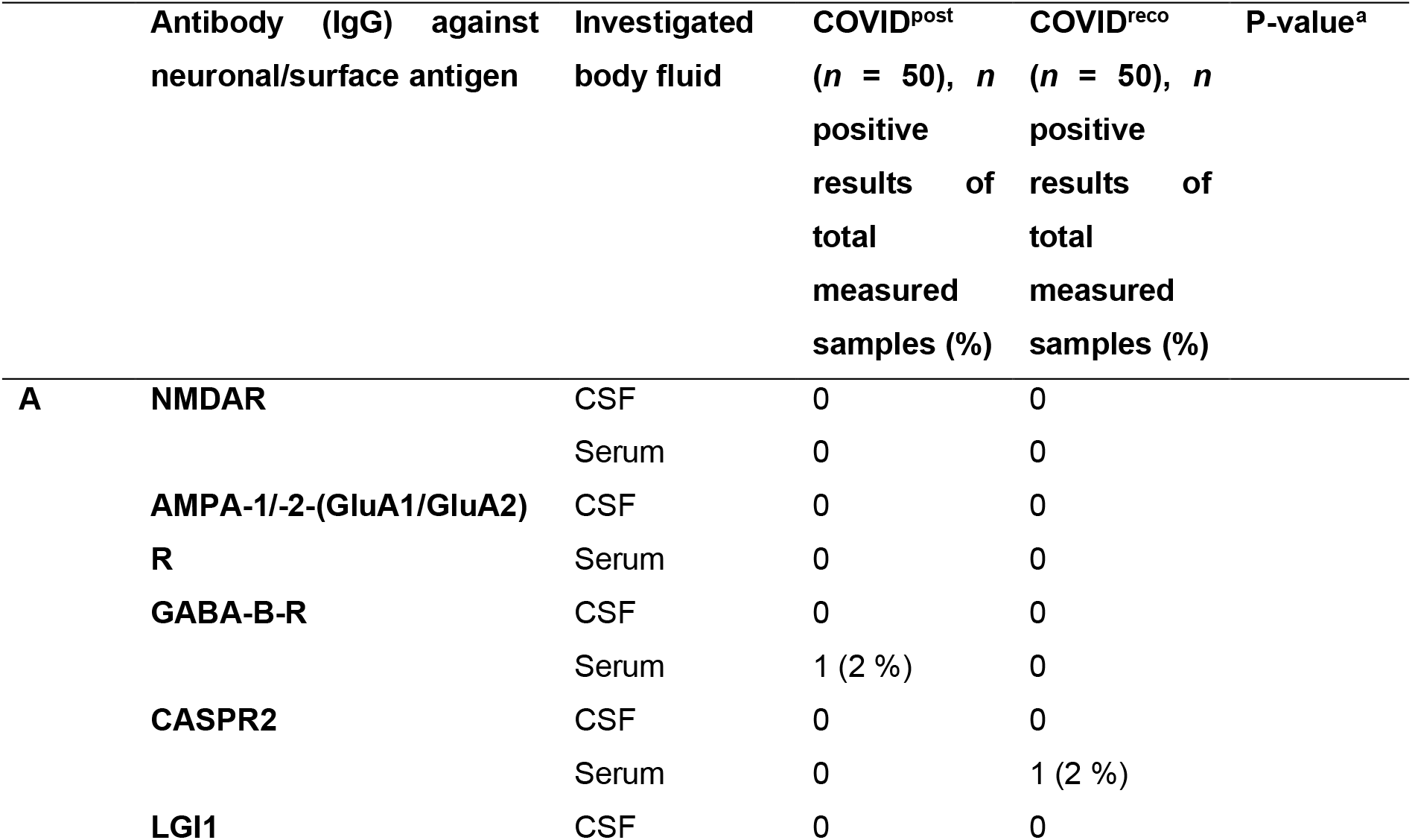
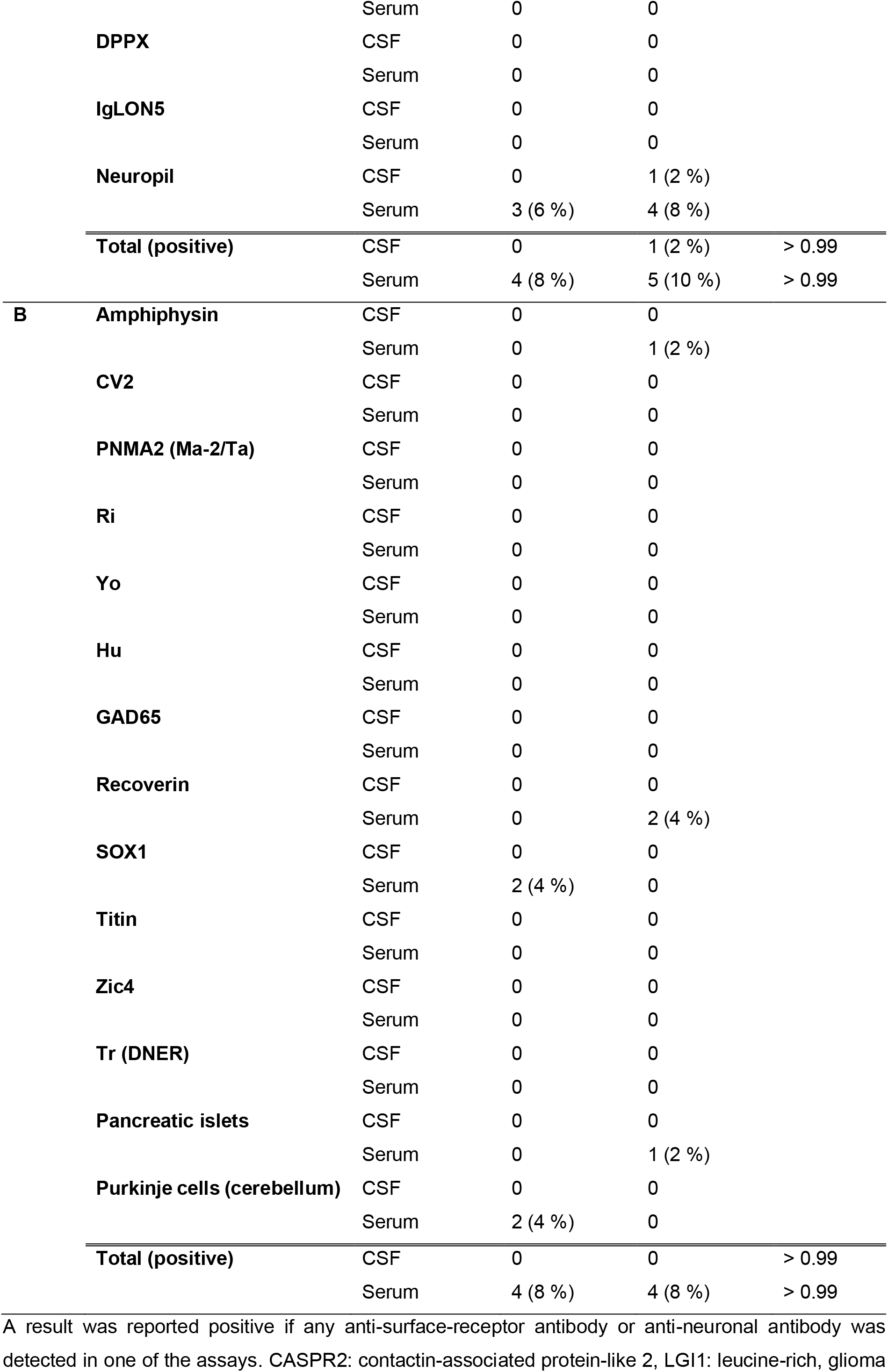
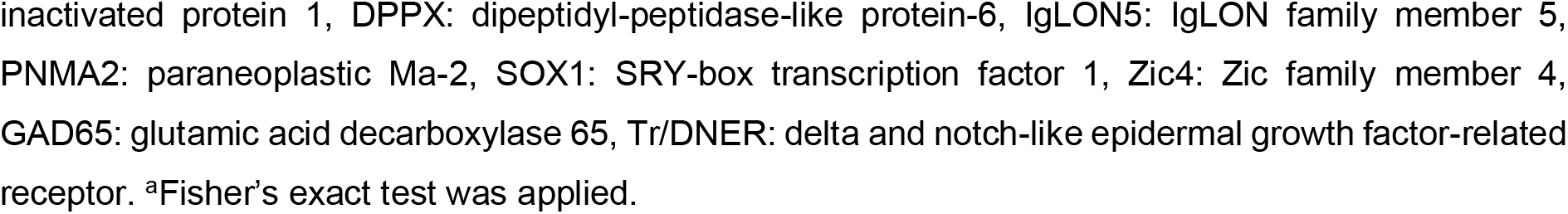
Findings of anti-neuronal autoantibody testing using commercial cell- and tissue-based assays in the COVID^reco^ and COVID^post^ cohorts.

Positive antineuronal antibody staining patterns, including reactivity against intracellular antigens in the line blot and IIFT, were detected in five COVID^post^ patients (4/50; 8 %) and four recovered individuals (4/50; 8 %) with no significant difference (*p* = 0.99). No corresponding antineuronal antibody reactivity was observed in the CSF. Among participants with positive antineuronal antibody findings, reactivity was directed against SOX1 (*n* = 2, 1+, only in line blot, not in IIFT), amphiphysin (*n* = 1, 3+, not in IIFT, but AAB against neuropil in serum), and recoverin (*n* = 2, 1+ and 2+), while IIFT additionally revealed reactivity against Purkinje cells (*n* = 2, no cerebellar symptoms), and pancreatic islet cells (*n* = 1, only in IIFT).

### Associations between neuropsychological outcomes and SARS-CoV-2-specific antibody measures

Across all regression models, neither SARS-CoV-2-specific antibody indices, Q_IgG_ values, nor serum or CSF SARS-CoV-2-specific IgG concentrations showed association with global cognition, total fatigue, cognitive fatigue, or motor fatigue (all *p* > 0.05; Supplementary Table S5). Likewise, no interactions between group status and any SARS-CoV-2-specific antibody measure were observed. Only the COVID^post^ cohort was associated with lower global cognition function and higher scores in all fatigue outcomes (η²p = 0.21 - 0.81, all *p* < 0.001).

## Discussion

Despite a distinct clinical phenotype of PCS regarding cognitive impairment and cognitive and motor fatigue, we found no evidence of CSF-specific alterations in this prospective single-center study distinguishing patients with PCS (COVID^post^) from individuals who had fully recovered following acute COVID-19 (COVID^reco^). Routine CSF analyses, SARS-CoV-2-specific antibody measurements, and antineuronal intracellular as well as neuronal surface antibody testing yielded largely comparable results between cohorts. Furthermore, SARS-CoV-2-specific antibody measures showed no association with cognitive function or fatigue.

In line with previous rRT-PCR analyses of CSF obtained from patients with acute COVID-19 and PCS, SARS-CoV-2 RNA was not detected in either CSF or blood samples in our cohort of 100 participants.^7–11,13,20,33^ Viral dissemination in the CSF or peripheral blood of patients with PCS seems not to be associated with the development of post-COVID-19 symptoms after an acute infection in our cohort. However, there are limited data on reservoirs of SARS-CoV-2 RNA in CSF and brain sections according to autopsy studies detecting viral RNA even 230 days after acute COVID-19.^17,18,59,60^ Furthermore, intrathecally produced SARS-CoV-2 antibodies were not detected which is in line with two previously published studies which also investigated patients with PCS and mainly neuropsychiatric symptoms,^10,20^ but in contrast to Bernard-Valnet et al. who detected elevated CoV-AIs in 62.5% of acute COVID-19 cases (5/8).^8^ Notably, SARS-CoV-2-specific IgG antibodies were detectable in almost all CSF samples of our cohort. However, given the CSF/serum CoV-AIs not exceeding the upper limit of 1.5 in almost all cases except two and the absence of severe blood-CSF barrier dysfunction, this finding is unlikely to reflect cohort-specific intrathecal antibody synthesis.^50^ According to the Reiber model, serum-derived immunoglobulins continuously enter the CSF through physiological blood-CSF exchange processes depending on their hydrodynamic radius and molecular weight.^47^ Therefore, in individuals with circulating SARS-CoV-2-specific antibodies, low concentrations of these antibodies are expected to be detectable in CSF even in the absence of CNS-directed immune activation, as observed in our both cohorts. Consequently, the widespread detection of SARS-CoV-2-specific IgG antibodies in CSF most likely represents physiological transfer from the systemic circulation rather than evidence of a compartmentalized intrathecal immune response. Further supporting a systemic origin of SARS-CoV-2-specific antibodies in CSF, these IgG concentrations in CSF correlated positively with both serum antibody levels and Q_Alb_. The interpretation of intrathecal SARS-CoV-2-specific antibody synthesis in 62.5% reported by Bernard-Valnet et al. should be considered with caution.^8^ A substantial proportion of the calculated SARS-CoV-2-specific AIs of the whole cohort in the study by Bernard-Valnet et al. fell below 0.7, a range generally regarded as physiologically implausible within the Reiber framework and therefore not reliably interpretable.^51^ Given that SARS-CoV-2-specific IgG concentrations in CSF were frequently close to the lower limit of detection, minor analytical variations may have disproportionately influenced quotient calculations and the resulting AIs. Consequently, the robustness of the reported AI results may be limited, and the evidence for intrathecal antibody synthesis appears less conclusive than suggested by the authors.^8^ Virus-specific intrathecal antibody synthesis may persist for several years after recovery from a CNS infection, as described for HSV1 encephalitis, neurosyphilis, and neuroborreliosis.^61–63^ Mean time from COVID-19 to LP were 1.85 years in our COVID^post^ cohort. Given that pathogen-specific intrathecal antibody synthesis is known to persist for years following CNS infections, the absence of elevated SARS-CoV-2 AIs in our cohort makes it unlikely that intrathecal SARS-CoV-2-specific antibody synthesis had occurred during the acute phase of infection. However, it should be noted that our cohort consisted almost exclusively of individuals who had experienced mild acute COVID-19, with 96% of the COVID^post^ cohort and 100% of recovered participants classified as WHO CPS stage 1 and nobody exhibited neurological manifestations during the acute infection. These findings are further supported by a large multicentric study of 127 patients with acute COVID-19, which likewise failed to demonstrate evidence of SARS-CoV-2-specific CNS immune activation.^33^ Despite no evidence of intrathecal immune activation in PCS, subtle compartment-specific immune alterations cannot be fully excluded. Investigations of peripheral blood mononuclear cells (PBMCs) have provided evidence for persistent immune activation in PCS. Following stimulation of circulating CD4+ and CD8+ T lymphocytes, patients with PCS exhibited increased intracellular levels of IL-2, IL-4, and IL-6 compared with convalescent controls.^64^ Evidence for intrathecal immune activation has primarily been derived from studies in acute COVID-19. In CSF, compartmentalized upregulation of genes related to IL-1β and IL-12 signaling was observed in Th1 and Th2 CD4+ T cells, accompanied by enhanced expression of canonical activation pathways in CD8+ T cells, collectively indicating a prominent T-cell-mediated immune response within the CNS.^65^ In addition, B-cell-related pathways appeared to be involved in the central immune response to SARS-CoV-2.^65^ Distinct plasma cell clusters were identified in the CSF, and in a murine model expressing the human angiotensin-converting enzyme 2 (ACE2) receptor, intracranial SARS-CoV-2 infection resulted in virus replication restricted to the brain and the exclusive detection of anti-spike antibodies in the CSF.^65^ Studies investigating routine CSF findings in PCS are limited.^10,19,20,24^ However, these studies lacked a neuropsychologically well-described control cohort of individuals who had fully recovered from COVID-19.

Blood-CSF barrier dysfunction has previously been reported as a common finding in studies investigating CSF alterations during acute COVID-19 with neurological manifestations.^7,8,11,33^ In the largest CSF study conducted to date in acute COVID-19, Q_Alb_ was elevated in 50 % of cases (58/116) and remained increased in 48 % and 46 % of patients even beyond 14 and 30 days after neurological symptom onset, respectively.^33^ Greene et al. found in a small cohort of 21 patients (11 with PCS and 10 fully recovered) that patients reporting brain fog and having cognitive deficits according to the Montreal Cognitive Assessment (MoCA) exhibited increased leaky blood vessels predominantly in both temporal lobes and in the left and right frontal cortex in dynamic contrast-enhanced MRI.^26^ By contrast, blood-CSF barrier dysfunction was detected in only 13% of participants (11/84) in a PCS cohort, indicating a substantially lower prevalence than that reported in studies of acute COVID-19.^19^ We found dysfunction of the blood-CSF barrier in 20% of the COVID^post^ cohort and 22% of the COVID^reco^ patients which is comparable to the prevalence of individuals with no evidence of a neurological disease (15 %).^66^

Beyond intrathecal virus-specific immune responses and blood-CSF barrier dysfunction, CNS-directed autoimmunity has also been discussed as a potential pathophysiological mechanism underlying PCS.^25^ This hypothesis is partly based on observations from HSV1 encephalitis, where secondary autoimmune responses against neuronal antigens, mainly NMDAR, can occur following the acute phase.^67^ In contrast to Boesl et al.^19^ who found serum, but not CSF, antibodies against myelin in 18% (12/68) of the PCS cohort, we could not detect any PCS-specific findings regarding anti-neuronal/surface antibodies in line with a former study.^10^ However, the clinical significance of antineuronal antibody reactivity detected exclusively in serum remains controversial, particularly in patients presenting with non-specific neurological symptoms such as cognitive impairment and fatigue.^68^ As a limitation, antibody titers are not given and the authors did not specify the exact assays used (ELISA, line blot, cell-based assays, and immunohistochemistry) making comparisons difficult.^19^ Boesl et al. also discussed the already reported high titers of myelin antibodies in healthy individuals.^19^ Furthermore, a recent study detected specific antibodies against myelin, Ma2/Ta, GAD65, Yo, and NMDAR (all serum) next to various undetermined epitopes using indirect immunofluorescence on unfixed murine brain sections (serum and CSF).^21^ CSF-autoantibodies correlated significantly with cognitive impairment according to MoCA.^21^ However, in contrast to our study, a representative control cohort was not included and antibody titers were not displayed.^21^ Moreover, a recent review reported low-frequency serum positivity for NMDAR (25/3065 [0.8 %]) and CASPR2 (5/2238 [0.2 %]) antibodies even in healthy controls.^69^ Given the fact that antibody positivity was typically detected in either the line blot or the IIFT rather than consistently across both assays and the absence of corresponding CSF reactivity in most cases of our cohort, these findings should be interpreted with caution and may represent either false-positive or unspecific findings. A few positive antineuronal staining patterns, including neuropil reactivity, were detected at comparable frequencies in the COVID^post^ and COVID^reco^ cohorts and were not associated with corresponding CSF antibody findings. In the absence of cohort-specific enrichment or evidence of intrathecal antibody production, these serum reactivities should be interpreted with caution.

### Strengths and limitations

A major strength of this study is the comprehensive clinical, neuropsychological, and neuroimaging characterization of both the PCS and fully recovered cohorts, combined with routinely available CSF and serum diagnostics, enhancing the clinical applicability and reproducibility of our findings. Additionally, PCS was diagnosed according to the established WHO Delphi consensus criteria and after an extensive interdisciplinary exclusion workup to increase confidence that the observed findings are attributable to PCS rather than competing diagnoses.^34^

Although the study may have been underpowered to detect small effect sizes (70% power to detect a medium effect size (Cohen’s *d* = 0.5) at a two-sided α of 0.05), the absence of significant group differences was not accompanied by borderline non-significant findings or consistent directional trends, thereby reinforcing the overall negative findings. Furthermore, neuropsychological examination was missing in 14 participants of the COVID^reco^ cohort. Missing data were handled automatically by the statistical software packages GraphPad Prism and R according to the requirements of the respective analyses. The interpretation of the observed blood-CSF barrier dysfunction in both cohorts is further limited by the absence of pre-infection or acute-phase CSF analyses. Therefore, it cannot be determined whether these findings were pre-existing. Another limitation concerns the COVID^reco^ cohort. Recovery from SARS-CoV-2 infection was assessed based on medical history rather than objective confirmation. Furthermore, except for 10 healthy controls, participants in the COVID^reco^ cohort presented to our hospital because of the symptoms and medical conditions described in the Methods section. Therefore, although these individuals did not fulfill the diagnostic criteria for post-COVID-19 syndrome, the cohort may not fully represent an entirely asymptomatic population.

## Conclusion

In conclusion, this prospective study provides no evidence that persistent neurological symptoms after COVID-19 are associated with ongoing adaptive CNS immune activation, intrathecal SARS-CoV-2-specific antibody synthesis, or disease-specific neuronal autoimmunity. The inclusion of a carefully phenotyped COVID-19 recovered comparison cohort strengthens the conclusion that the observed CSF findings do not seem to reflect mechanisms specific to post-COVID syndrome. Although patients with PCS exhibited substantial cognitive impairment and severe fatigue, these symptoms were not associated with routine CSF inflammatory markers or SARS-CoV-2-specific humoral immune responses. Our findings therefore argue against routine CSF diagnostics as a source of disease-specific biomarkers in unselected PCS patients and instead support future research focusing on alternative mechanisms such as neuroglial dysfunction, altered brain network connectivity, vascular dysfunction, and persistent systemic immune dysregulation.

## Data availability statement

Anonymous data is available from qualified investigators upon reasonable request.

## Acknowledgements

We want to thank Refika Aksamija, Joleene Holm, Nicole Renske, Tatiana Simak, Vera Lehmensiek, Sandra Hübsch, Dagmar Schattauer, Alice Beer, Stephanie Becker, and Martina Leis from the Laboratory of Cerebrospinal Fluid Diagnostics and Clinical Neurochemistry, Biobank, and Autoimmune Laboratory of the Neurological Department of the University of Ulm (Germany).

