## Supplementary for "Persistent Neurological Symptoms After COVID-19 Lack Evidence of Adaptive CNS Immune Activation"

### Supplementary Material

**Table S1.** Additional demographical and clinical data of the COVID$^{post}$ and COVID$^{reco}$ cohort.

| Parameter | COVID$^{post}$ ($n$ = 50) | COVID$^{reco}$ ($n$ = 36) | Test statistics[a] |
|---|---|---|---|
| **WHO Clinical Progression Scale[1]** | | | $p$ = 0.51[#] |
| **1 (mild) (x/total), %** | 48/50; 96 % | 36/36; 100 % | |
| **2 (moderate) (x/total), %** | 2/50; 4 % | 0/36 | |
| **3 (severe) (x/total), %** | 0/50 | 0/36 | |
| **Psychiatric lifetime diagnosis** | | | $p$ = 0.004 |

| | | | |
|---|---|---|---|
| **Anxiety disorder (x/total), %** | 3/50; 6 % | 2/36; 6 % | |
| **Depression (x/total), %** | 16/50; 32 % | 2/36; 6 % | |
| **PTSD (x/total), %** | 2/50; 4 % | 0/36 | |
| **None (x/total), %** | 29/50; 58 % | 32/36; 89 % | |
| **Fatigue (FSMC)** | | | $p < 0.001$ |
| **None (x/total), %** | 0/50 | 23/36; 64 % | |
| **Mild (x/total), %** | 0/50 | 10/36; 28 % | |
| **Moderate (x/total), %** | 2/50; 4 % | 2/36; 6 % | |
| **Severe (x/total), %** | 48/50; 96 % | 1/36; 3 % | |
| **SDMT (mean ± SD, z-score)** | -0.96 ± 1.15 | -0.19 ± 0.74 | $t = -3.38$, $p = 0.0014$ |

Only patients from the COVID$^{reco}$ cohort with complete neuropsychological data were included in the analyses presented in this table ($n$ = 36). [a]P-values < 0.05 were set as statistically significant. Fisher's exact test and Pearson's $\chi^2$ test were used for categorical variables, where appropriate. Continuous variables were analyzed using a bootstrapped Yuen's test (trimmed means, $tr$ = 0.2). [#]Odds ratios were not reported because one cell contained a zero count, resulting in an undefined estimate due to division by zero. PTSD: post-traumatic stress disorder, FSMC: fatigue scale for motor and cognitive functions, SDMT: symbol digit modalities test.

**Table S2.** Post-infectious symptoms of the COVID$^{post}$ cohort.

| **Symptom** | **COVID$^{post}$** ($n$ = 50) |
|---|---|
| **Cognitive deficits (x/total), %** | 50/50; 100 % |
| **Fatigue, incl. PEM (x/total), %** | 50/50; 100 % |

| | |
|---|---|
| **Headache (x/total), %** | 25/50; 50 % |
| **Dizziness (x/total), %** | 9/50; 18 % |
| **Myalgia (x/total), %** | 24/50; 48 % |
| **Arthralgia (x/total), %** | 15/50; 30 % |
| **Sleep disorder (x/total), %** | 31/50; 62 % |
| **Sensory disturbance (x/total), %** | 13/50; 26 % |
| **Tremor (x/total), %** | 1/50; 2 % |
| **Chronic pain syndrome (x/total), %** | 11/50; 22 % |
| **Exertional dyspnea (x/total), %** | 12/50; 24.0% |
| **Resting dyspnea (x/total), %** | 4/50; 8 % |
| **Postural tachycardia (HR** ≥ 100 bpm) **(x/total), %** | 17/50; 34 % |
| **Tinnitus (x/total), %** | 4/50; 8 % |
| **Visual disturbance (x/total), %** | 4/50; 8 % |
| **Hyposmia/Anosmia (x/total), %** | 4/50; 8 % |
| **Dysgeusia (x/total), %** | 2/50; 4 % |
| **Gait/movement disorder (x/total), %** | 2/50; 4 % |
| **Fever/subfebrile (x/total), %** | 7/50; 14 % |
| **Altered bowl habits (diarrhea/ constipation) (x/total), %** | 10/50; 20 % |
| **Abdominal pain (x/total), %** | 9/50; 18% |
| **Nocturnal dry cough (x/total), %** | 1/50; 2 % |
| **Flu-like symptoms/recurrent URTI (x/total), %** | 16/50; 32 % |
| **Hair loss (x/total), %** | 1/50; 2 % |
| **Photophobia/phonophobia (x/total), %** | 12/50; 24% |

Values are given as positive absolute and relative frequency from total $n = 50$. PEM: post-exertional malaise, HR: heart rate, bpm: beats per minute, URTI: upper respiratory tract infection.

**Table S3.** Additional CSF routine findings of the COVID$^{post}$ and COVID$^{reco}$ cohort.

| Parameter | COVID$^{post}$ ($n$ = 50) | COVID$^{reco}$ ($n$ = 50) | Test statistics[a] |
|---|---|---|---|
| **Blood-CSF barrier dysfunction acc. to $Q_{Alb}$** | | | $p = 0.59$[#] |
| **None (x/total), %** | 40/50; 80.0% | 39/50; 78.0% | |
| **Slight (x/total), %** | 10/50; 20 % | 10/50; 20 % | |
| **Moderate (x/total), %** | 0/50 | 1/50; 2 % | |
| **OCB pattern** | | | $p = 0.33$[#] |
| **1 (x/total), %** | 27/50; 54 % | 24/50; 48 % | |
| **2 (x/total), %** | 2/50; 4 % | 1/50; 2 % | |
| **3 (x/total), %** | 4/50; 8 % | 1/50; 2 % | |
| **4 (x/total), %** | 17/50; 34 % | 24/50; 48 % | |

Categorical variables are presented as positive absolute and relative frequencies from total $n$ of each cohort. [a] Fisher's exact test was used for the categorical variables. P-values < 0.05 were set as statistically significant. [#]Odds ratios were not reported because one cell contained a zero count, resulting in an undefined estimate due to division by zero. OCB: oligoclonal IgG bands.

**Table S4.** Demographical, clinical, neuropsychological, radiographic, and CSF analytical findings of patients with elevated AI[a] of both cohorts as indicated.

| No. Fig. 4 | Cohort | AI[a] | Demographics | PCS symptoms | Neuropsychological findings | CSF findings | MRI findings | Comorbidities |
|---|---|---|---|---|---|---|---|---|

| | | | | | | | | |
|---|---|---|---|---|---|---|---|---|
| P1 | **COVID$^{post}$** | CoV 4.1 | f, 1.49 y since COVID-19, WHO CPS 1 | Tremor, cognitive deficits, fatigue, exertional dyspnea | Global cognition[b]: -0.39, total fatigue[c]: 102, cognitive fatigue[c]: 56, motor fatigue[c]: 46 | WBCC 1/µl, lactate 1.45 mmol/l, $Q_{Alb}$ 4.2 x $10^{-3}$, OCB pattern 4, anti-neuronal/surface AB negative | Unremarkable | None |
| P5 | **COVID$^{reco}$** | CoV 17.7 | f, 1.72 y since COVID-19, WHO CPS 1 | None | Global cognition[b]: -0.01, total fatigue[c]: 20, cognitive fatigue[c]: 10, motor fatigue[c]: 10 | WBCC 1/µl, lactate 1.40 mmol/l, $Q_{Alb}$ 5.1 x $10^{-3}$, OCB pattern 1, anti-neuronal/surface AB negative | Empty sella | Idiopathic intracranial hypertension |
| P2 | **COVID$^{post}$** | M 4.0, R 1.7 | f, 4.15 y since COVID-19, WHO CPS 1 | Cognitive deficits, fatigue, myalgia, sleep disorder, paresthesia, exertional/resting dyspnea, postural tachycardia | Global cognition[b]: -3.53, total fatigue[c]: 96, cognitive fatigue[c]: 49, motor fatigue[c]: 47 | WBCC 1/µl, lactate 1.47 mmol/l, $Q_{Alb}$ 5.1 x $10^{-3}$, OCB pattern 3, anti-neuronal/surface AB negative | Subcortical white matter lesions | None |
| P6 | **COVID$^{reco}$** | M 2.0 | f, 1.37 y since COVID-19, WHO CPS 1 | None | Global cognition[b]: 0.74, total fatigue[c]: 23, cognitive fatigue[c]: 11, motor fatigue[c]: 12 | WBCC 1/µl, lactate 1.75 mmol/l, $Q_{Alb}$ 9.4 x $10^{-3}$, OCB pattern 3, anti- | Subcortical white matter lesions | Arterial Hypertension |

| | | | | | | | | |
|---|---|---|---|---|---|---|---|---|
| | | | | | | neuronal/surface AB negative | | |
| P4 | **COVID$^{post}$** | HSV1 2.5 | f, 2.0 y since COVID-19, WHO CPS 1 | Cognitive deficits, fatigue, myalgia, sleep disorder, exertional dyspnea, altered bowel habits, abdominal pain | Global cognition[b]: 0.6, total fatigue[c]: 83, cognitive fatigue[c]: 37, motor fatigue[c]: 46 | WBCC 6/µl, lactate 1.41 mmol/l, $Q_{Alb}$ 6.2 x $10^{-3}$, OCB pattern 2, anti-neuronal/surface AB negative, multiplex PCR for neurotropic viruses neg. | Unremarkable | None |
| P3 | **COVID$^{post}$** | Z 6.7 | f, 1.54 y since COVID-19, WHO CPS 1 | Cognitive deficits, fatigue, headache, myalgia, sleep disorder, exertional dyspnea | Global cognition[b]: -3.27, total fatigue[c]: 85, cognitive fatigue[c]: 41, motor fatigue[c]: 44 | WBCC 5/µl, lactate 1.65 mmol/l, $Q_{Alb}$ 5.1 x $10^{-3}$, OCB pattern 2, anti-neuronal/surface AB negative | Unremarkable | None |
| P7 | **COVID$^{reco}$** | Z 1.7 | m, 1.82 y since COVID-19, WHO CPS 1 | None | Global cognition[b]: -3.27, total fatigue[c]: 60, cognitive fatigue[c]: 32, motor fatigue[c]: 28 | WBCC 3/µl, lactate 1.37 mmol/l, $Q_{Alb}$ 5.6 x $10^{-3}$, OCB pattern 4, anti-neuronal/surface AB negative | Unremarkable | None |

| | | | | | | | | |
|---|---|---|---|---|---|---|---|---|
| P8 | **COVID**$^{reco}$ | CMV 3.0 | m | None | - | WBCC 1/µl, lactate 1.48 mmol/l, $Q_{Alb}$ 2.9 x $10^{-3}$, OCB pattern 4, anti-neuronal/surface AB negative | Unremarkable | Idiopathic intracranial hypertension |
| P9 | **COVID**$^{reco}$ | CMV 4.8 | m, 0.45 y since COVID-19, WHO CPS 1 | None | Global cognition[b]: -3.27, total fatigue[c]: 51, cognitive fatigue[c]: 26, motor fatigue[c]: 25 | WBCC 1/µl, lactate 1.18 mmol/l, $Q_{Alb}$ 5.1 x $10^{-3}$, OCB pattern 4, anti-neuronal/surface AB negative | Unremarkable | None |

[a]AI: virus-specific CSF/serum antibody index, [b]Composite score, [c]: FSMC: Fatigue scale for motor and cognitive functions, AB: antibody, PCS: post-COVID-19 syndrome, y: year, WHO CPS: WHO Clinical Progression Scale[1]

**Table S5.** Association of specific CSF/serum findings with global cognition and fatigue - moderation by group.

| Outcome | Parameter | β [95%CI] | Statistic | *p* | $\eta^2p$ [95%CI] |
|---|---|---|---|---|---|
| **Global cognition (standardized)** | Cohort[a] | -0.42 [-0.59 - -0.24] | $t(82) = -4.82$ | < 0.001 | 0.22 [0.08 - 0.37] |
| | $Q_{IgG\_total}$ (standardized) | -0.02 [-0.22 - 0.17] | $t(82) = -0.23$ | 0.816 | 0 [0 - 0.05] |
| | Cohort[a] × $Q_{IgG\_total}$ (standardized) | -0.08 [-0.28 - 0.11] | $t(82) = -0.84$ | 0.404 | 0.01 [0 - 0.09] |
| **Global cognition (standardized)** | Cohort[a] | -0.41 [-83.13 - 82.31] | $t(73) = -0.01$ | 0.992 | 0 [0] |

| Outcome | Parameter | β [95%CI] | Statistic | *p* | η²p [95%CI] |
|---|---|---|---|---|---|
| | CoV-AI (standardized) | 0.3 [-769.13 - 769.74] | *t*(73) = 0 | 0.999 | 0 [0] |
| | Cohort[a] × CoV-AI (standardized) | 0.3 [-769.14 - 769.74] | *t*(73) = 0 | 0.999 | 0 [0] |
| **Global cognition (standardized)** | Cohort[a] | -0.42 [-0.6 - -0.23] | *t*(77) = -4.48 | < 0.001 | 0.21 [0.07 - 0.36] |
| | [Serum SARS-CoV-2-specific IgG] (standardized) | 0.03 [-0.13 - 0.2] | *t*(77) = 0.38 | 0.708 | 0 [0 - 0.06] |
| | Cohort[a] × [Serum SARS-CoV-2-specific IgG] (standardized) | 0.03 [-0.14 - 0.2] | *t*(77) = 0.36 | 0.722 | 0 [0 - 0.06] |
| **Global cognition (standardized)** | Cohort[a] | -0.39 [-10.9 - 10.11] | *t*(73) = -0.07 | 0.941 | 0 [0] |
| | [CSF SARS-CoV-2-specific IgG] (standardized) | 0.46 [-86.13 - 87.04] | *t*(73) = 0.01 | 0.992 | 0 [0] |
| | Cohort[a] × [CSF SARS-CoV-2-specific IgG] (standardized) | 0.45 [-86.13 - 87.04] | *t*(73) = 0.01 | 0.992 | 0 [0] |
| **FSMC (total) (standardized)** | Cohort[a] | 0.92 [0.82 - 1.01] | *t*(82) = 18.48 | < 0.001 | 0.81 [0.73 - 0.85] |
| | $Q_{IgG\_total}$ (standardized) | 0.04 [-0.06 - 0.14] | *t*(82) = 0.82 | 0.416 | 0.01 [0 - 0.09] |
| | Cohort[a] × $Q_{IgG\_total}$ (standardized) | -0.05 [-0.15 - 0.05] | *t*(82) = -1.03 | 0.305 | 0.01 [0 - 0.1] |
| **FSMC (total) (standardized)** | Cohort[a] | 0.91 [-30.16 - 31.98] | *t*(73) = 0.06 | 0.954 | 0 [0] |
| | CoV-AI (standardized) | 0.15 [-288.93 - 289.23] | *t*(73) = 0 | 0.999 | 0 [0] |
| | Cohort[a] × CoV-AI (standardized) | 0.21 [-288.87 - 289.29] | *t*(73) = 0 | 0.999 | 0 [0] |
| **FSMC (total) (standardized)** | Cohort[a] | 0.9 [0.79 - 1.01] | *t*(77) = 16.45 | < 0.001 | 0.78 [0.69 - 0.83] |
| | [Serum SARS-CoV-2-specific IgG] (standardized) | 0 [-0.22 - 0.22] | *t*(77) = -0.01 | 0.996 | 0 [0] |

| Outcome | Parameter | β [95%CI] | Statistic | *p* | η²p [95%CI] |
|---|---|---|---|---|---|
| | Cohort[a] × [Serum SARS-CoV-2-specific IgG] (standardized) | -0.02 [-0.24 - 0.2] | $t(77) = -0.15$ | 0.877 | 0 [0 - 0.04] |
| **FSMC (total) (standardized)** | Cohort[a] | 0.89 [-13.71 - 15.49] | $t(73) = 0.12$ | 0.904 | 0 [0 - 0.04] |
| | [CSF SARS-CoV-2-specific IgG] (standardized) | -0.05 [-120.4 - 120.29] | $t(73) = 0$ | 0.999 | 0 [0] |
| | Cohort[a] × [CSF SARS-CoV-2-specific IgG] (standardized) | 0 [-120.34 - 120.35] | $t(73) = 0$ | 1.000 | 0 [0] |
| **FSMC (cognitive) (standardized)** | Cohort[a] | 0.9 [0.79 - 1.01] | $t(82) = 16.42$ | < 0.001 | 0.77 [0.68 - 0.82] |
| | $Q_{IgG\_total}$ (standardized) | 0.01 [-0.09 - 0.11] | $t(82) = 0.16$ | 0.874 | 0 [0 - 0.04] |
| | Cohort[a] × $Q_{IgG\_total}$ (standardized) | -0.04 [-0.14 - 0.06] | $t(82) = -0.75$ | 0.457 | 0.01 [0 - 0.08] |
| **FSMC (cognitive) (standardized)** | Cohort[a] | 0.89 [-13.39 - 15.18] | $t(73) = 0.12$ | 0.901 | 0 [0 - 0.04] |
| | CoV-AI (standardized) | 0.2 [-132.54 - 132.95] | $t(73) = 0$ | 0.998 | 0 [0] |
| | Cohort[a] × CoV-AI (standardized) | 0.27 [-132.48 - 133.01] | $t(73) = 0$ | 0.997 | 0 [0] |
| **FSMC (cognitive) (standardized)** | Cohort[a] | 0.88 [0.76 - 1] | $t(77) = 14.33$ | < 0.001 | 0.73 [0.62 - 0.8] |
| | [Serum SARS-CoV-2-specific IgG] (standardized) | 0.02 [-0.24 - 0.28] | $t(77) = 0.14$ | 0.889 | 0 [0 - 0.04] |
| | Cohort[a] × [Serum SARS-CoV-2-specific IgG] (standardized) | -0.02 [-0.28 - 0.24] | $t(77) = -0.15$ | 0.878 | 0 [0 - 0.04] |
| **FSMC (cognitive) (standardized)** | Cohort[a] | 0.87 [-16.88 - 18.62] | $t(73) = 0.1$ | 0.923 | 0 [0 - 0.03] |
| | [CSF SARS-CoV-2-specific IgG] (standardized) | -0.05 [-146.34 - 146.24] | $t(73) = 0$ | 0.999 | 0 [0] |

| Outcome | Parameter | β [95%CI] | Statistic | *p* | η²p [95%CI] |
|---|---|---|---|---|---|
| | Cohort[a] × [CSF SARS-CoV-2-specific IgG] (standardized) | 0.01 [-146.29 - 146.3] | $t(73) = 0$ | 1.000 | 0 [0] |
| **FSMC (motor) (standardized)** | Cohort[a] | 0.91 [0.82 - 1.01] | $t(82) = 18.91$ | < 0.001 | 0.81 [0.74 - 0.86] |
| | $Q_{IgG\_total}$ (standardized) | 0.08 [-0.03 - 0.18] | $t(82) = 1.45$ | 0.150 | 0.03 [0 - 0.12] |
| | Cohort[a] × $Q_{IgG\_total}$ (standardized) | -0.06 [-0.16 - 0.05] | $t(82) = -1.11$ | 0.272 | 0.01 [0 - 0.1] |
| **FSMC (motor) (standardized)** | Cohort[a] | 0.9 [-46.65 - 48.44] | $t(73) = 0.04$ | 0.970 | 0 [0] |
| | CoV-AI (standardized) | 0.09 [-442.23 - 442.42] | $t(73) = 0$ | 1.000 | 0 [0] |
| | Cohort[a] × CoV-AI (standardized) | 0.15 [-442.18 - 442.47] | $t(73) = 0$ | 0.999 | 0 [0] |
| **FSMC (motor) (standardized)** | Cohort[a] | 0.9 [0.8 - 1.01] | $t(77) = 17.07$ | < 0.001 | 0.79 [0.71 - 0.84] |
| | [Serum SARS-CoV-2-specific IgG] (standardized) | -0.02 [-0.21 - 0.18] | $t(77) = -0.2$ | 0.843 | 0 [0 - 0.05] |
| | Cohort[a] × [Serum SARS-CoV-2-specific IgG] (standardized) | -0.01 [-0.21 - 0.18] | $t(77) = -0.14$ | 0.887 | 0 [0 - 0.04] |
| **FSMC (motor) (standardized)** | Cohort[a] | 0.88 [-10.32 - 12.09] | $t(73) = 0.16$ | 0.876 | 0 [0 - 0.04] |
| | [CSF SARS-CoV-2-specific IgG] (standardized) | -0.04 [-92.39 - 92.3] | $t(73) = 0$ | 0.999 | 0 [0] |
| | Cohort[a] × [CSF SARS-CoV-2-specific IgG] (standardized) | 0.01 [-92.33 - 92.35] | $t(73) = 0$ | 1.000 | 0 [0] |

Linear regression results with heteroscedasticity-robust standard errors and bootstrapped confidence intervals. Continuous variables were z-standardized and the grouping variable was effect-coded. [a]$COVID^{post}$ and $COVID^{reco}$, FSMC: Fatigue scale for motor and cognitive functions, $Q_{IgG\_total}$: CSF $IgG_{total}$/serum $IgG_{total}$, CoV-AI: SARS-CoV-2-specific CSF/serum IgG antibody index
